# Beyond episodic early warning systems: a continuous clinical alert system for early detection of in-hospital deterioration

**DOI:** 10.1101/2025.05.20.25327940

**Authors:** Michael R. Scheid, Beth Friedmann, Michael Oppenheim, Jamie S. Hirsch, Theodoros P. Zanos

## Abstract

Efficient patient monitoring on the medical-surgical wards is crucial to prevent significant in-hospital adverse events. Standard episodic inpatient assessment of vital signs can potentially miss changes in health status and delay recognition of elevated risk. To reduce the likelihood of this delayed recognition of risk, we developed a wearable-based deep learning model, using only 9 inputs, to identify the onset of deterioration earlier than traditional early warning systems. We showed this model could generalize to produce clinical alerts ahead of a broad class of significant adverse clinical outcomes, including rapid response team (RRT) interventions, unplanned intensive care unit (ICU) transfers, intubations, cardiac arrests, and in-hospital deaths. Using data from 888 adult non-ICU inpatient visits in four hospitals in New York and employing two different clinical grade wearable biosensors (4-8% data missingness, excluding SpO2), as part of a quality initiative, we trained a recurrent neural network (RNN) to predict both MEWS alerts and adverse clinical outcomes. Using multiple stages of validation, we showed in our retrospective, time-sequence duration optimized, prospective validation the RNN model was able to predict both periods of elevated MEWS scores (ROC AUC 0.89 +/- 0.3, PR AUC 0.58 +/- 0.14) and adverse clinical outcomes (accuracy: 81.8% on 11 events) up to an average of 17 hours in advance. Our results show that our wearable based RNN alert system outperforms traditional episodic clinical support tools in detecting early onset of inpatient deterioration; enabling timely interventions that can improve outcomes and reduce hospital costs for patients in early stages of deterioration.

## Introduction

Up to 5% of patients admitted to non–critical care inpatient units develop deterioration requiring escalation of care and transfer to a critical care setting^1^. Delayed recognition of this critical illness is associated with increased morbidity and mortality^2–8^. Since the current standard of non-intensive care unit (ICU) care relies on intermittent vital signs measured every four to eight hours, the intervening period between measurements can result in missed warning signs of deterioration. Even in inpatient settings with continuous monitoring (CM) solutions in place, such as ICUs, step-down units, or telemetry units, many monitoring systems use simple thresholds or cutoffs that often result in a predictable cascade of events: over-alerting leading to hospital staff alert-fatigue^9^; dismissal of useful alerts; delayed recognition of critical illness^10^; and, ultimately, worse outcomes^2–7^. In conjunction with more discriminating alerting systems, continuous CM has the potential to facilitate timely and accurate early identification of deterioration on inpatient units.

While a large body of research and early warning scores (EWS) have focused on facilitating early identification of the deteriorating patient, newer wearable technologies that continuously monitor vital signs can help detect vital sign abnormalities sooner and thereby reduce provider workload through automated capture^11,12^. Enabling patient monitoring while minimizing staff exposure to infected patients, remote monitoring options through wearables received increased attention during the coronavirus disease 2019 (COVID-19) pandemic. These devices also expanded the ability of health systems to monitor the increased number of acute, high-risk, and/or rapidly deteriorating patients^13^, and some studies have explored the ability of these devices to improve outcomes^11,12,14–16^. In one of the few studies with many patients^15^ (over 7000), CM on wards was associated with significant decreases in total length of hospital stay, ICU days, and code blue rates. Other studies have reported lower rapid response team (RRT) rates^17^, decreases in ICU transfers^16,18^, reduced lengths of stay^16,19,20^, and lower readmission rates^20^. Overall, however, the clinical and operational impact of continuous monitoring is uncertain given a limited number of studies, mostly with small sample sizes^11,16^. Furthermore, these studies rely on standard alerting thresholds or existing deterioration models.

Solely based on episodically collected electronic health record (EHR) data, many models have been developed to predict risk of clinical outcomes for hospitalized patients^21–26^. Importantly, in most clinical predictive models, inference is anchored and performed at a certain point of the patient trajectory—for example, upon hospitalization^27^, transfer to the ICU^28–30^, or first or last clinical observations^31^. While similar approaches have been used in studies utilizing CM data^28–35^, CM-based deterioration algorithms can also be continuous with constantly updating inferences. Despite existing studies on continuously updating ICU-based mortality models using CM data^32,34,35^, no studies for medical-surgical ward patients have been published. Interpretation of multimodal continuous biometric data in this context opens the possibility for new methods to detect significant physiologic patterns to better predict the early onset of deterioration. The few studies that collected CM data focused on validation of the CM measurements, evaluation of the feasibility of deploying CM devices^36^, or correlations of existing clinical scores with various clinical outcomes (such as EWS)^12,37–40^. Predictive models developed from CM data that have been reported in select small (i.e., < 100) outpatient populations demonstrate the feasibility of this approach^41,42^. Further, it has been shown that deep learning models achieve the best performance on continuous monitoring data compared to many existing modelling approaches, hence our selection of a deep learning model^57^. Thus, models that use larger numbers of CM monitored inpatients to evaluate whether they improve detection of inpatient deterioration need to be developed and evaluated.

In this study, we piloted the use of CM devices in four of our health system hospitals and collected CM data from 888 non-ICU inpatients (i.e., 2,897 patient days). We validated the correlation between CM and standard vital sign measurements, assessed the feasibility and benefits of using these data to generate clinical alerts, and developed a deep-learning model using CM data to identify patient deterioration up to 24 hours before an EHR-based Modified Early Warning Score (MEWS) clinical alert. We collected CM data using two chest-worn wearable devices and validated the deep-learning model in three stages using three unique datasets: data held out for testing from the device used to train the model (Retrospective Device #1), prospective data from Device #1 at a separate hospital (Prospective Device #1), and data from a second wearable device (Alternate Device #2) used on a different patient population at a different hospital to function as an alternate device validation. To establish both the accuracy and specificity of a real-time implementation of this CM-based inpatient deterioration detection algorithm, we simulated deployment of this algorithm by generating a stream of continuous inferences at all timepoints where patients were wearing a wearable device. We then measured the performance of this algorithm in detecting deterioration, as well as the rate of false alerts, at all timepoints in which valid CM data was available.

## Methods

### Data sources, patient population and eligibility criteria

The study design and protocol was approved by Northwell’s Institutional Review Board (study #: 23-0534-350CD). We obtained all EHR data collected for 888 hospitalized patients that received the CM devices between March 2020 and November 2022 across four hospitals within the Northwell Health system. We collected the CM data using two chest worn wearable devices, Vital Connect Vital Patch (Device #1) and Biobeat Chest Monitor (Alternate Device #2). The episodic data were collected from the enterprise EHR (Sunrise Clinical Manager, Altera [formerly Allscripts]) and included demographics (e.g., age and body mass index [BMI]) as well as manually monitored vital signs (i.e., heart rate [HR], systolic blood pressure [SBP], respiratory rate [RR], skin and body temperature, and oxygen saturation [SpO2]) and the continuously monitored vital signs (i.e., HR, RR, temperature, and SpO2). It should be noted that the wearable device provided both a skin temperature measurement and body measurement, which reflects core body temperature. Upon placement of the patch, the body temperature is calibrated by the nurse. For the wearable data validation we used the body temperature measurement from the device. Wearable device data was provided monthly by the device manufacturers. Table 1a provides a brief summary of the data source for each of the continuous monitoring and demographic variables:

**Table 1a.**
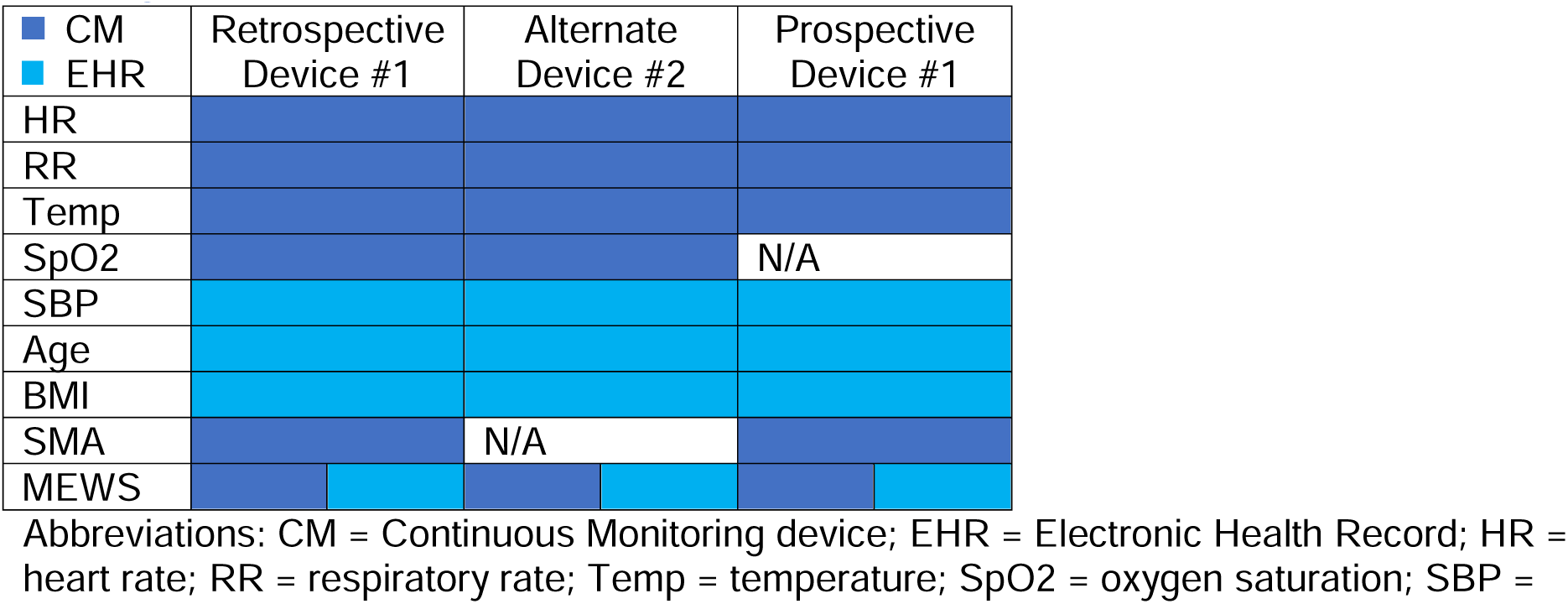
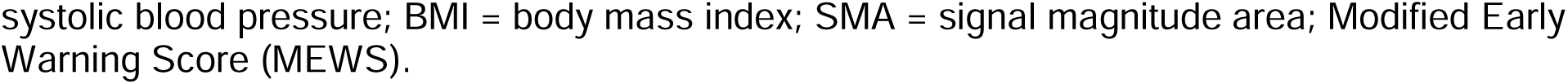
summarized data sources for each of the continuous monitoring and demographics variables.

While our retrospective analysis was approved by IRB as a research study, the patching of patients was performed as a quality initiative, that augmented the standard of care for these patients. The patients that were patched were selected based often on the best clinical judgement of the staff in each of the wards where the wearable devices were deployed. In practice, this meant the nurse overseeing the patient would often decide whether to have a wearable device placed on the patient or not based on their assessment of how at risk the patient was for decompensating.

### Study design

We sought to develop and evaluate, in a real-world clinical environment, a wearable clinical alert system (Figure 1). Our goals were to assess whether these devices were a viable clinical alternative for monitoring on the medical wards, whether increased frequency of vital signs provide an advantage over episodic monitoring, and whether the data acquired combined with a machine learning approach can yield an accurate predictive algorithm. We followed the steps outlined in Figure 1, diagram A, by aligning (step one) and visualizing (step two) differences between continuously monitored vitals and episodically monitored vitals using Bland-Altman plots (see green panel in Figure 1). To examine any advantages CM provides over episodic monitoring (see yellow panel in Figure 1, diagram A), we compared the numbers and timing of clinical alerts across measurement modalities. Finally, we used the clinical alerts as labels to train a model and predict periods of clinical deterioration further into the future than current early warning systems (see blue panel in Figure 1, diagram A).

**Figure 1.**
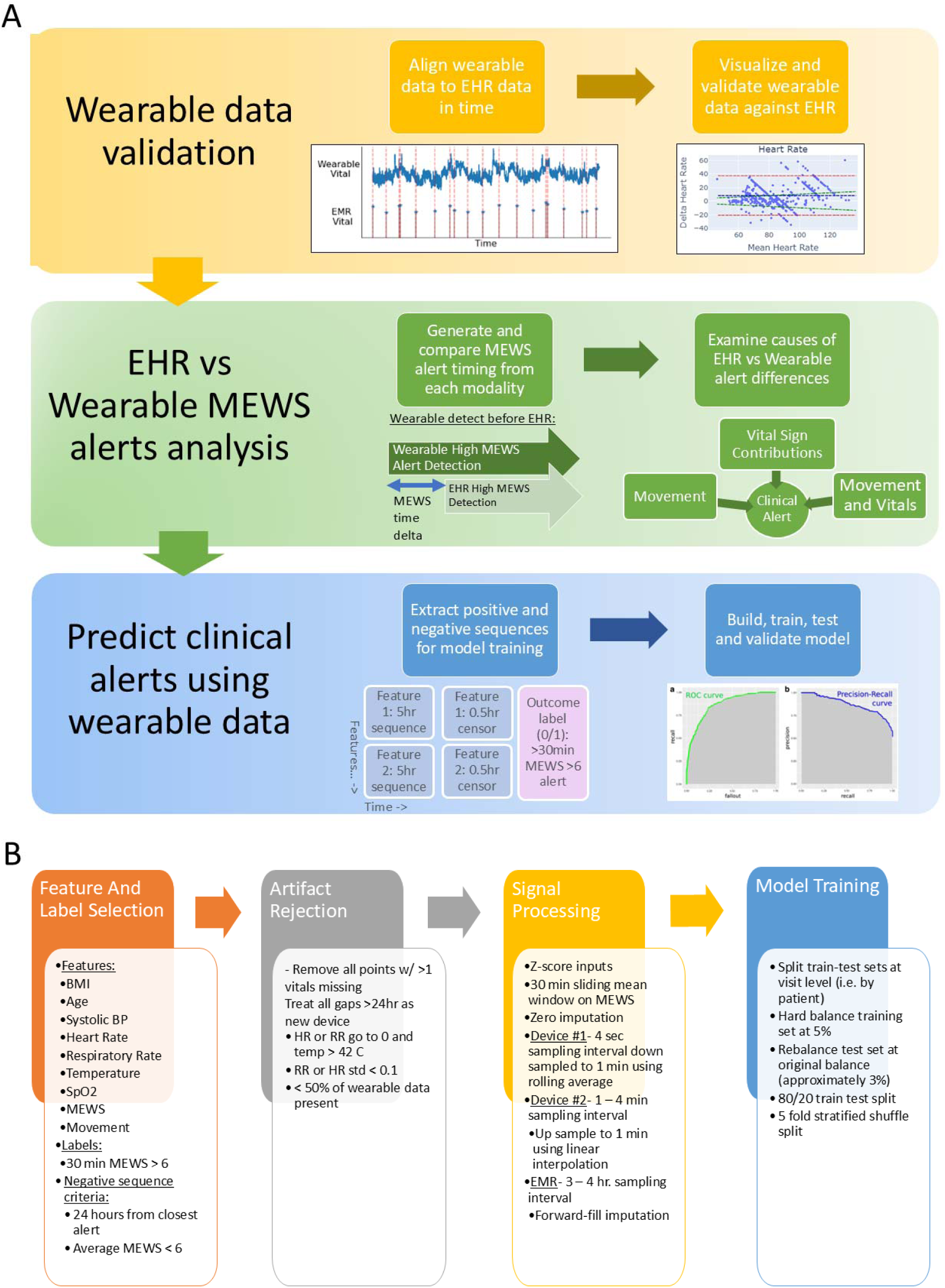
Flow diagrams of the validation stages for a wearable-based alert system and, processing and modelling steps. (Diagram A) The three stages of clinical validation for a wearable-based alert system (Yellow Panel) Wearable data accuracy validation ensures the data is of sufficient accuracy to use for clinical decision making and this to end we align (1st step) and visualize differences (2nd step) using Bland-Altman plots, that visualize the difference between the two modalities (y-axis) vs their mean (x-axis). (Green Panel) Comparison of clinical alerts across measurement modalities demonstrates whether a wearable device provides an advantage over episodic monitoring, where we compare the numbers and timing of alerts for each modality and calculated the sources of these differences. (Blue Panel) Wearable clinical alert modeling and validation showing the feature and label construction and performance of this algorithm. (Diagram B) Feature and label selection, artifact rejection, signal processing and model training and testing steps followed to build a wearable device based clinical alert system.

### Wearable data evaluation

All data processing, analysis and modeling was performed in Python using the Pandas, Numpy, Scikit-learn, Tensorflow, and Keras packages. Visualizations were created using the Matplotlib and Plotly packages.

We compared the vital sign measurements by synchronizing the CM vital signs with the manually acquired vital signs. To synchronize the two data sources, the wearable data was converted from UNIX time to human-readable time, shifted to account for both daylight-savings and time zone differences and then resampled to align to EHR data timestamps. The EHR data was organized chronologically by patient ID number and timestamps and resampled to align to the closest minute with the wearable data.

Using this time-aligned data, we constructed Bland-Altman plots, which visualize the agreement between two measurement modalities by plotting the difference (i.e., *Δ*) between the time-aligned signal from the two modalities (*Δ* = *Signal* 1 (*Wearable*) – *Signal* 2 (*EHR*)) on the y-axis against their mean 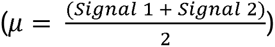 on the x-axis. The greater the agreement between the two modalities, the closer each of the points on the plot is to the horizontal line, where *Δ* = 0. For visualization purposes, we plotted the 95% confidence intervals (i.e., red dotted lines), mean delta (i.e., blue dotted line) and either the 5% or 10% error line (*Δ* = µ * 0.1) for each of the Bland-Altman plots. We also calculated the percentage of measurements that fell within the 5% or 10% error line. In subsequent analyses, we only used records where at least 2/3 (i.e., 67%) of a value for a given patient were within a 10% error for HR and a 5% error for temperature and SpO2 from the EHR values. This criterion was not applied to respiratory rate because of the well-documented digit preference among healthcare workers when manually measuring the respiratory rate^43^.

### Clinical alert analysis

To assess the benefits of using wearable data to produce more and earlier clinical alerts, we compared the volume, timing, and duration of alerts that arose from continuous versus episodic monitoring. We used the MEWS score^44^ as the basis for our clinical alerts for a number of reasons: it is derived from measurements acquired across all inpatients; includes a number of vitals that are measured by the wearable device; and is already implemented as part of the standard of care in many hospitals and health systems, including Northwell Health, where it is calculated automatically within the EHR and clinicians are alerted when defined thresholds are exceeded (Supplementary table 3).

We generated two distinct sets of MEWS scores—one derived from continuous wearable vitals and the other from episodic vitals—which resulted in two sets of alerts. The MEWS calculation is based upon vital sign measurements given by the equation below:

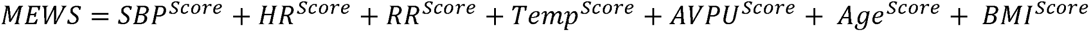

Depending on the range of each item, it is given a score ranging from 0 to 3 (Supplementary Table 2), and the final score is tallied (range 0-15), with higher values indicating greater risk of clinical decompensation. It should be noted that this a commonly used score, not only in our system, but in health systems around the world, as an early warning system. Based upon Northwell Health system protocol (Supplementary table 3) and expert consensus among our clinical collaborators, we defined a meaningful clinical alert as a MEWS > 6 for 30 minutes or more. We quantified the total number of alerts, onset timing, and duration for each alert. To understand the underlying cause of the differences between CM- and EHR-based clinical alerts, we measured the contribution of each vital sign to the MEWS score from each modality at the time of alert onset.

### Movement analysis

To rule out the possibility that motion-related events caused temporary changes in vital signs and thereby triggered alerts unrelated to early signs of deterioration, we analyzed biomarkers with respect to movement during each alert. Movement was captured by the device with a three-axis accelerometer and represented as the signal magnitude area (SMA), the sum of a one-second average of accelerations in all three axes^45^. The alerts and their corresponding average SMA values, ranging from two minutes preceding the alert onset to the end of the alert, were grouped into three classes: “stationary,” “activities of daily living (ADL),” and “strenuous,” based on manufacturer determined SMA cutoff values^45^ (Supplementary table 1).

### Predictive deterioration modeling

#### Artifact rejection, signal processing, imputation and model selection

Figure 1, Panel B illustrates the artifact rejection and signal processing steps applied to the data. We implemented artifact rejection to remove segments of data where the measurements assumed physiologically implausible values either due to lost connectivity or low battery issues with the device (i.e., HR or RR were zero and the temperature was above 50□C). In order to determine the optimal window length for predicting clinical alerts 0.5 to 24 hours in the future, we performed a grid search of window lengths from 1 to 12 hours at 1-hour increments and determined 5 hours to be the optimal length based on retrospective model performance. For the purposes of predicting 0.5 to 24 hours into the future, sufficient data of 5.5 hours (i.e., 5-hour sequence + 0.5-hour censor) preceding the clinical alert label were needed. We removed the segments and labels where < 50% of data was present within the 5-hour sequence.

The two devices that were used to continuously monitor vital signs had different sampling rates and were therefore processed differently to make them compatible with the same deep learning model. Device #1 had a four-second sampling interval and was down sampled to a one-minute interval using a one-minute-wide rolling mean window with a one-minute stride. Alternate Device #2 was sampled variably with a one- to fifteen-minute sampling interval and was up-sampled to one minute using linear interpolation. The EHR data was sampled with a three- to five-hour sampling interval and was up-sampled to one minute using forward-filling imputation. We z-scored the input features to bring them to the same scale for faster model convergence. Whenever continuous data was missing, we used 30 minutes forward filling and imputed with zeros beyond 30’. For the prospective Device #1, SpO2 was not available so the variables was imputed with all zeroes for the final model.

#### Model training and testing

We trained a recurrent neural network (RNN) with long-short-term-memory (LSTM) units model that incorporated age, BMI, SBP, and continuously monitored vital signs (HR, RR, temperature, SpO2, MEWS value, and motion signals) to predict in-hospital deterioration with a 0.5-24 hour time horizon using a 5 hour sequence (Figure 2). We constructed the clinical alert labels detailed previously, based on smoothed MEWS values using a 30-minute-wide rolling average window.

**Figure 2.**
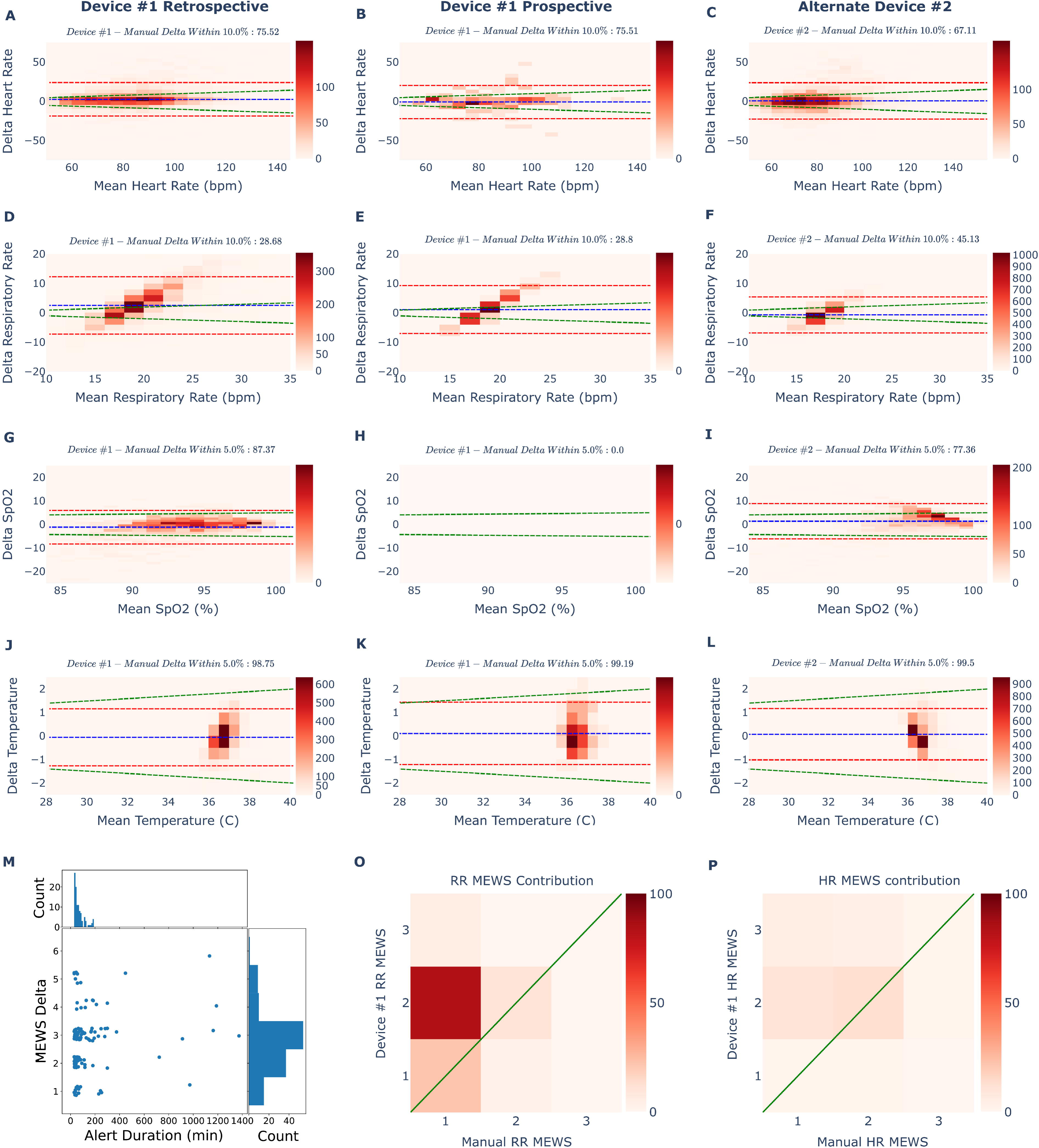
(Panels A-L) Bland Altman plots of the comparison between vital measurement from the CM devices and EHR vitals, for the retrospective, prospective and alternate device datasets. (Panels M-P) Features of elevated MEWS score alerts, detected by CM device and missed by EHR, and contribution of CM vitals in their generation. Dotted red lines indicate the 95% confidence intervals of the deltas between wearable measurements and EHR measurements. Dotted blue lines indicate the mean delta between wearable and EHR measurements. Green dotted lines indicate a 10% deviation of the wearable measurement from the EHR measurement at that mean value. Sub-panel titles detail the percentage of vital measurements that fall within the 10% deviation line (green dotted line). Bland Altman plots of (Panel A - C) Device #1 retrospective HR, Device #1 Prospective HR and Alternate Device #2 HR; (Panel D - E) Device #1 Retrospective RR, Device #1 Prospective RR and Alternate Device #2 RR; (Panel G - I) Device #1 Retrospective SpO2, Device #1 Prospective SpO2 (Device #1 Prospective dataset did not have SpO2) and Alternate Device #2 SpO2. (Panel J - L) Device #1 Retrospective temperature, Device #1 Prospective temperature, and Alternate Device #2 temperature. Two sets of wearable clinical alerts were generated post-hoc using vitals from Device #1 and EHR, respectively. Differences in the resulting alert onsets and magnitudes were compared between these two sets. (Panel M) Scatterplot and marginal distributions of the duration of wearable clinical alerts (MEWS >6) that were detected by the wearable but missed by the EHR, along with the difference in magnitude of the MEWS score, between wearable vs EHR derived MEWS, at alert onset. (Panel O, P) Heatmaps of MEWS contribution of RR. The wearable MEWS score was plotted on the y-axis and the EHR MEWS score on the x-axis. The diagonal green line represents equal scores given to both the wearable- and EHR-based vitals. Off diagonal values indicate that one modality was given a higher score than the others. We focused on respiratory rate and heart rate measurements, since both the temperature and SpO2 measurements were congruent between EHR and CM values. (Panel O) and HR (Panel P) from the wearable device (y-axis) against the EHR (x-axis, labelled ‘Manual’).

In order to select a model to test on clinical alerts and hard outcomes we performed hyperparameter optimization across the following parameters: number of LSTM layers, sequence length, time horizon, and censor length. The model selected showed superior performance across the various validation steps performed in this study.

Class imbalance was addressed by first randomly removing samples from the negative majority class and oversampling the positive class until we achieve a balance of 95%/5% negative/positive classes of the training set, instead of the original 99.7%/0.3% class balance. We evaluated the predictive performance of this model in three stages: retrospective, prospective, and alternate Device validations. In the first retrospective validation stage, we validated the model using 5-fold stratified shuffle split cross-validation at the patient level, i.e., no patients in the training set were included in the test set. This resulted in an approximately 80%/20% train/test split. We then compared predictive performance across all folds, using common model validation metrics (ROC AUC, PR AUC) and averaged the resulting ROC and PR curves. After the initial validation stage, we then tested the model in two additional validation stages using two unique datasets: 1) data collected prospectively from Device #1, i.e. entirely after Device #1’s retrospective data used to train the model, at a separate hospital and from a different patient cohort, and 2) data from a second wearable device used used at a separate hospital and on an entirely different patient population to function as an alternate device test set. In each subsequent stage of validation, i.e. “Prospective” and “Alternate Device” validations, the model that was trained exclusively on Device #1’s retrospective dataset was then tested on each of these pure validation datasets. Neither data from prospective Device #1 data nor Alternate Device #2 was used to optimize, hyperparameter tune or train the retrospectively trained model in anyway. These datasets, and their accompanying validation results, therefore functioned as two true unique test sets for the retrospectively optimized and trained model.

These three validation stages contrasted performance of the LSTM RNN model against logistic regression (LR) using two concurrently collected datasets: 1) CM data from the wearable device and EHR data collected alongside that wearable data (EHR).

#### Model Calibration

Calibration curves were constructed for the LSTM RNN model across the three stages of validation (Supplementary Figure. 5). Isotonic regression recalibration was performed on the calibration curves for each stage of validation. Brier scores (BS) were also calculated on all calibration curves. All RNN performance metrics were recalculated using the recalibrated model (Figure. 3, Panels A-F)

**Figure 3.**
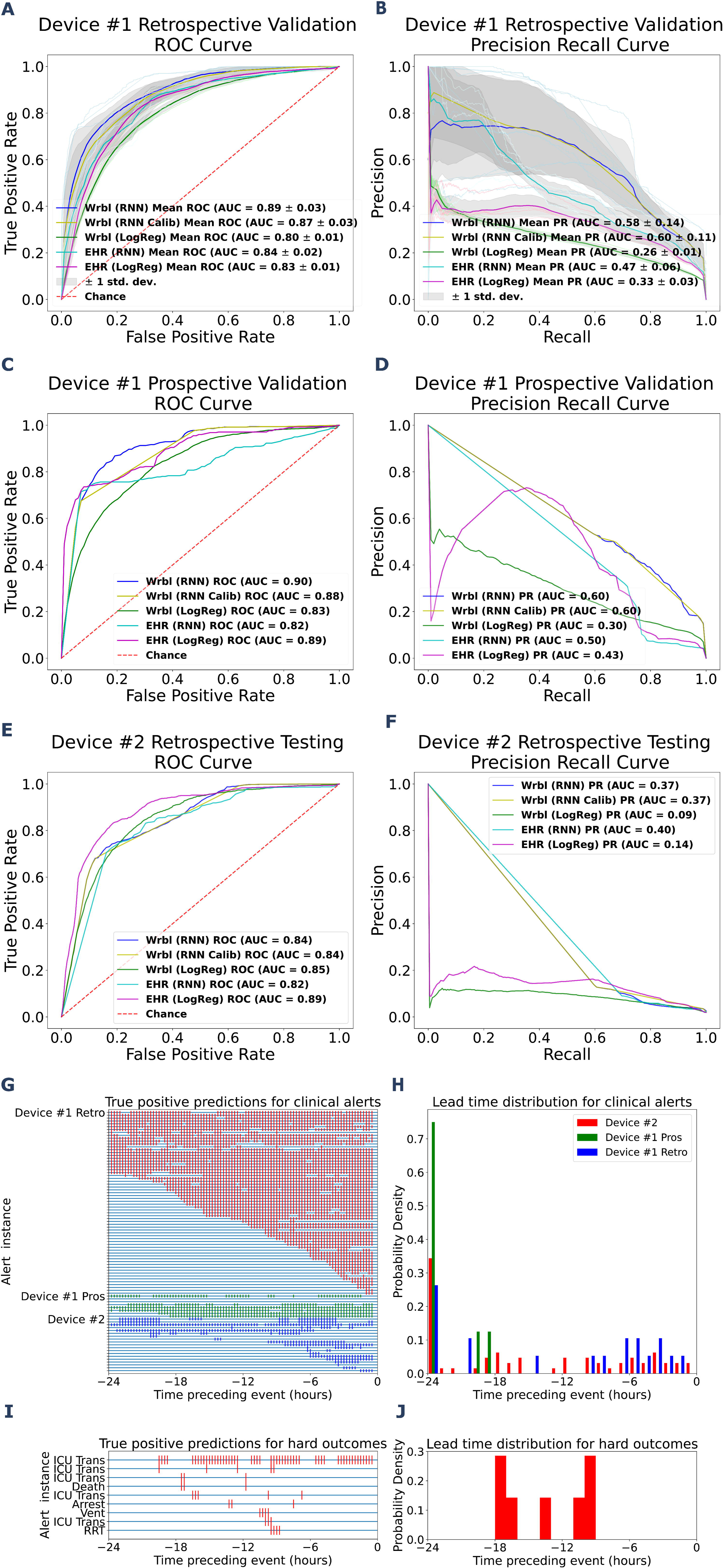
(Panels A-J) Wearable (Wrbl) based Recurrent Neural Network (RNN), Recalibrated Wearable (Wrbl) based Recurrent Neural Network (RNN Calib), Logistic Regression (LogReg) and EHR based model performances illustrated by receiver operator characteristic (ROC) and precision-recall (PR) curves for twenty-four types of validation. (Panels G-J) Lead time analysis of wearable based RNN model illustrates how far in advance model can predict both clinical alerts and hard outcomes. ROC and PR curves for predicting periods 30mins or greater of continuously elevated MEWS (MEWS > 6) 0.5-24 hours into the future. AUCs for each curve are listed in the figure legend. (Panel A, B) Retrospective RNN, Recalibrated RNN and LR model performances in predicting an elevated MEWS using randomly shuffled Device #1 data, and concurrently collected EHR data, which was held out for testing. Performance of each model, for each dataset, across folds is plotted (lighter shade). Mean performances are plotted (darker shade) with one standard deviation of performances across folds also plotted (grey). Chance performance of a binary classifier is plotted (dotted red line). (Panel C, D) RNN, Recalibrated RNN and LR Model performances in predicting an elevated MEWS using prospective data from the same device (Device #1 Prospective) and concurrently collected EHR data, which were both held out entirely for testing. (Panel E, F) RNN, Recalibrated RNN and LR Model performances in predicting an elevated MEWS using data from another device (Alternate Device #2), and concurrently collected EHR data, which were both held out entirely for testing. Lead times here are the timing of the first true positive preceding an alert. ‘ICU Trans’ = Unplanned ICU Transfer, ‘Arrest’ = Cardiac Arrest, ‘Vent’ = Ventilator, ‘RRT’ = Rapid response team call (Panel G) Raster plot visualization of timing of all true positive predictions made by RNN model (not recalibrated) with wearable data preceding a clinical alert (Time 0). The colors represent the various datasets--Red raster: Device #1 Retrospective dataset, green raster: Device #1 Prospective dataset, blue raster: Alternate Device #2 dataset. (Panel H) Probability density histogram of the lead times of the RNN model (not recalibrated) predictions for each alert. Colors correspond to those in the raster plot. (Panel I) Raster plot visualization of timing of all true positive predictions (not recalibrated) relative to hard outcomes (Time 0). (Panel J) Probability density histogram of the lead times of the RNN model (not recalibrated) predictions for each hard outcome.

### Testing wearable RNN model accuracy on hard outcomes

Using the RNN model trained to detect > 30-minute periods of elevated MEWS, we tested the model’s performance on prespecified “hard outcomes” with sufficient wearable data prior (i.e., at least 5.5 hours). In total 11 hard outcomes were found in the Retrospective Device #1 dataset with sufficient preceding wearable data in order to make predictions. We also ensured that the hard outcomes being predicted upon were at least 24 hours away from each other. To predict hard outcomes, we used the same features, processing steps, and testing procedure of the previous modeling steps with the exception that the label being predicted was one of five hard outcomes: RRT call, unplanned transfer to the ICU, intubation/mechanical ventilation, cardiac arrest, and death (expiration). Successful detections were counted if a true positive was generated within the 24 hours leading up to the outcome. The performance of the RNN algorithm in detecting hard outcomes was compared using CM data against EHR data alone. A comparison was also made using an elevated MEWS (> 6) as the predictor for the hard outcome.

### Lead time and sampling rate analysis

To characterize the timing of predictions relative to alerts, we looked at the lead times of predictions of the wearable-based RNN for each alert and hard outcome. Lead time was defined as the first true positive preceding alerts and hard outcomes.

To determine the effect of sampling rate on prediction performance, we varied the inter-sample timing of vital measurements and measured the effect on overall performance. Data were downsampled from the original inter-sample interval of one minute down to three hundred minutes. Performance was measured by ROC AUC and PR AUC at each inter-sample interval.

## Results

We piloted the use of two CM devices in four of our health system hospitals and collected CM data between March 2020 and November 2022 from 888 non-ICU inpatients over 2,897 patient days. Table 1 summarizes the demographics of the study population.

**Table 1:** Summarized demographics and statistics across all prediction cohorts.

|  | Total |  |  | Retrospective | Alternate Device | Prospective |
| --- | --- | --- | --- | --- | --- | --- |
|  | All | Stable | Deteriorated | All | All | All |
| Patients (n) | 888 | 871 | 17 | 315 | 413 | 160 |
| Devices (n) | 1302 | 1244 | 58 | 496 | 576 | 230 |
| Male (%) | 453 (51.2) | 445 (51.3) | 8.0 (53.3) | 162 (51.4) | 206 (49.9) | 85 (53.1) |
| Race/Ethnicities, n (%) |  |  |  |  |  |  |
| White | 648 (73.3) | 634 (73.1) | 14.0 (93.3) | 214 (67.9) | 328 (79.4) | 106 (66.2) |
| Multiracial | 94 (10.6) | 92 (10.6) | 2.0 (13.3) | 34 (10.8) | 36 (8.7) | 24 (15.0) |
| Black | 86 (9.7) | 85 (9.8) | 1.0 (6.7) | 32 (10.2) | 30 (7.3) | 24 (15.0) |
| Asian | 38 (4.3) | 38 (4.4) | 0 (0.0) | 26 (8.3) | 9 (2.2) | 3 (1.9) |
| Unknown/Other | 16 (2.1) | 18 (2.1) | 0 (0.0) | 9 (2.8) | 6 (1.5) | 6 (2.8) |
| Declined | 4 (0.5) | 4.0 (0.5) | 0 (0.0) | 0 (0.0) | 4 (1.0) | 0 (0.0) |
| Model Features, median [IQR] |  |  |  |  |  |  |
| Age [median IQR] | 69.3 [57.6, 78.0] | 69.3 (57.5 - 77.3) | 88.3 (86.5 - 90.5) | 68.0 [56.0, 78.0] | 70.0 [60.0, 79.0] | 70.0 [56.8, 77.0] |
| BMI [median IQR] | 27.0 [23.5 - 32.3] | 27.4 [23.5 - 32.3] | 30.4 (27.3 - 31.8) | 27.4 [23.7, 32.1] | 28.3 [23.9, 33.3] | 26.6 [23.0, 31.5] |
| SMA [median IQR] | 16.3 [8.9 - 17.7] | 16.3 [8.9 - 17.7] | 15.6 [9.1 - 16.3] | 16.3 [8.9 - 17.7] | 0.0 [0.0 - 0.0] | 0.0 [0.0 - 0.0] |
| sBP [median IQR] | 126.0 [109.3 - 137.7] | 125.8 [109.3 - 137.7] | 134.8 [120.3 - 147.8] | 125.2 [110.0 - 138.0] | 127.3 [112.0 - 140.0] | 125.5 [106.0 - 135.0] |
| HR [median IQR] | 79.8 [68.2 - 90.8] | 79.8 [68.1 - 90.8] | 79.8 [70.5 - 90.1] | 79.7 [67.3 - 90.7] | 75.7 [65.4 - 84.9] | 84.0 [71.8 - 96.7] |
| RR (mean SD) | 19.0 [16.5 - 21.3] | 19.0 [16.5 - 21.2] | 19.3 [17.3 - 21.2] | 21.0 [17.4 - 23.9] | 17.6 [16.0 - 19.1] | 18.5 [16.1 - 20.8] |
| Temp (mean SD) | 36.7 [36.3 - 36.8] | 36.7 [36.3 - 36.8] | 36.7 [36.3 - 37.1] | 36.7 [36.3 - 37.1] | 36.6 [36.4 - 36.7] | 36.5 [36.1 - 36.7] |
| SpO2 (mean SD) | 96.0 [95.0 - 97.9] | 96.5 [95.3 - 98.2] | 93.3 [91.1 - 96.3] | 93.0 [90.8 - 96.0] | 96.7 [95.4 - 99.0] | 0.0 [0.0 - 0.0] |
| MEWS (mean SD) | 3.0 [2.0 - 4.0] | 2.9 [2.0 - 3.9] | 5.6 [5.2 - 6.0] | 3.1 [2.2 - 4.0] | 3.0 [2.0 - 4.0] | 2.8 [1.8 - 4.0] |

Abbreviations: BMI = body mass index; HR = heart rate; IQR = interquartile range; Modified Early Warning Score (MEWS); RR = respiratory rate; SBP = systolic blood pressure; SD = standard deviation; SMA = signal magnitude area; SpO2 = oxygen saturation; Temp = temperature.

### Wearable data evaluation

Vital sign measurements from the wearable devices were evaluated by synchronizing and comparing with manually acquired vital signs logged in the EHR by constructing Bland-Altman plots (Figure 2). We only used wearable vitals for modeling when 67% of measured values were within 10% error from the EHR values.

The Retrospective and Prospective Device #1 and Alternate Device #2 HR respectively had 75%, 75%, and 67% of measurements falling within the 10% deviation line, indicating sufficient HR concordance for use in modeling (Figure 2A, 2B, 2C). Retrospective and Prospective Device #1 and Alternate Device #2 RR respectively had 29%, 29%, and 45% of measurements falling within the 10% deviation line (Figure 2D, 2E, 2F). As EHR RR is often inaccurately measured, our inclusion criteria were not applied for this vital sign. Retrospective Device #1 and Alternate Device #2 SpO2 respectively had 87% and 77% of measurements falling within the 5% deviation line (Figure 2G, 2I), while Prospective Device #1 did not include SpO2 measurements (Figure 2H). Retrospective and Prospective Device #1 and Alternate Device #2 temperature had 99% of all datasets’ and devices’ measurements fall within the 5% deviation line (Figure 2J, 2K, 3L). The threshold for acceptable agreement between the two measurements for both temperature and SpO2 readings was set at 5%, as ranges of normal values for these measurements are lower in clinical practice.

### Clinical alert, vital contribution, and movement analyses

Two sets of clinical alerts were generated post hoc using vitals from Device #1 and the EHR, respectively, with differences in the resulting alert volumes and magnitudes compared. The alert algorithm detected 140 clinical alerts, defined as a MEWS greater than 6 for 30 minutes or longer. The marginal histogram plot (Figure 2M) shows the number of elevated wearable-MEWS score events (not appearing in the EHR) plotted by delta in alert magnitude (wearable-MEWS – EHR-MEWS [x-axis]) against alert duration (y-axis). The CM devices detected 126 (or 9x) more alerts than manual monitoring (Figure 2M). Of all alerts, 90% were detected by Device #1 alone, while fewer than 3% were only detected in the EHR. For 7% of alerts that were detected by both, the wearable-based alerts preceded the EHR alerts by an average of 105 minutes.

The incongruity in alerts between wearable- and EHR-derived MEWS alerts was due to RR (Figure 2O) and HR (Figure 2P) measurements, contributing to higher MEWS scores and thus more alerts. In plotting the respective MEWS contributions of the vital signs from each modality against each other, RR and HR respectively contributed 30x and 10x as much to the MEWS scores as compared with other vital signs. For the 126 (i.e., 90%) of alerts that were missed by the EHR, accurate reporting of RR accounted for 89 (i.e., 71%) of these alerts, while elevated continuous HR measures accounted for 20 (i.e., 16%).

#### MWS

Since HR and RR can change significantly with patient movement, we sought to test whether movement-induced elevation of these two vitals might be contributing to elevated MEWS values (i.e., alerts) despite clinical and physiologic patient stability. We examined the values of the vitals (i.e., HR, RR) against the average SMA values over the duration of the detected alert and associated MEWS value. The lack of a correlation between increasing movement (i.e., SMA values) and increasing vital sign measurements (i.e., HR – R^2^ = 0.01, RR – R^2^ = 0.02) or MEWS values indicated that the alerts were generated independently of movement (Supplementary figures 1 and 2).

### Clinical alert modeling performance and validation

We trained an LSTM deep learning model using various demographic and continuously monitored vital sign features to predict in-hospital deterioration with a 24-hour time horizon using a 5-hour sequence. A logistic regression (LR) based model was also trained for comparison, and all models were compared to a dataset using EHR data alone without continuous device data. The predictive performance of these models was evaluated retrospectively (i.e., Device #1 held out data), prospectively (i.e., Device #1 separate hospital), and externally (i.e., Device #2 alternate device). Setting an operating point to achieve a positive predictive value of 0.2, we also examined the confusion matrix for each of the three validation steps (Table 2).

**Table 2:**
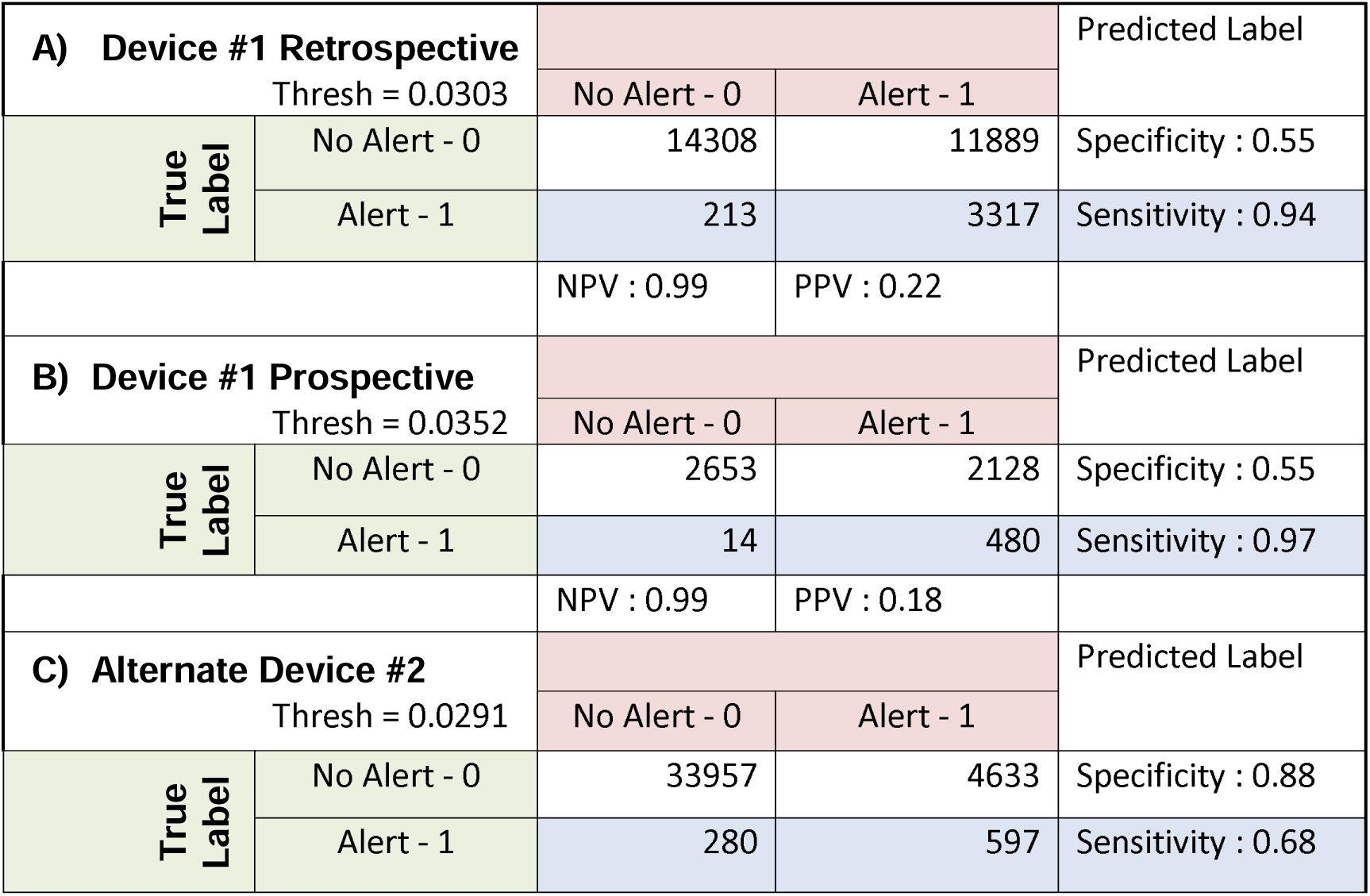
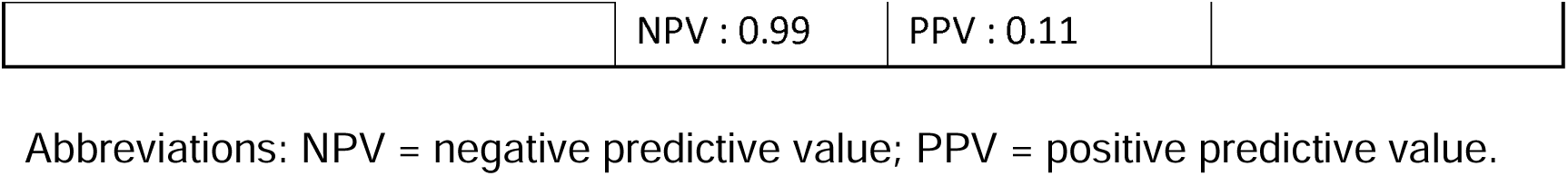
Confusion matrices for each stage of validation.

| **A) Device #1 Retrospective** |  |  |  | Predicted Label |
| --- | --- | --- | --- | --- |
| Thresh = 0.0303 |  | No Alert - 0 | Alert - 1 |  |
| True Label | No Alert - 0 | 14308 | 11889 | Specificity : 0.55 |
|  | Alert - 1 | 213 | 3317 | Sensitivity : 0.94 |
|  |  | NPV : 0.99 | PPV : 0.22 |  |
| **B) Device #1 Prospective** |  |  |  | Predicted Label |
| Thresh = 0.0352 |  | No Alert - 0 | Alert - 1 |  |
| True Label | No Alert - 0 | 2653 | 2128 | Specificity : 0.55 |
|  | Alert - 1 | 14 | 480 | Sensitivity : 0.97 |
|  |  | NPV : 0.99 | PPV : 0.18 |  |
| **C) Alternate Device #2** |  |  |  | Predicted Label |
| Thresh = 0.0291 |  | No Alert - 0 | Alert - 1 |  |
| True Label | No Alert - 0 | 33957 | 4633 | Specificity : 0.88 |
|  | Alert - 1 | 280 | 597 | Sensitivity : 0.68 |

|  |  |  |
| --- | --- | --- |
|  | NPV : 0.99 | PPV : 0.11 |

Abbreviations: NPV = negative predictive value; PPV = positive predictive value.

#### Retrospective validation on Device #1 data

Using data held out for testing from Device #1, we validated the RNN-based model to predict the occurrence of CM-based clinical alerts. The RNN model performance on this retrospective dataset (blue trace) had mean +/- SD ROC AUC 0.89 +/- 0.03 and PR AUC 0.58 +/- 0.14 as compared to the LR model (mean ROC AUC 0.80 +/- 0.01 and PR AUC 0.26 +/- 0.01, green trace—Figure 3A and 3B and Table 2). The RNN-based model had worse performance when using EHR data alone (turquoise trace—Figures 5A and 5B) versus device data with mean ROC AUC 0.84 +/- 0.02 and PR AUC of 0.47 +/- 0.06 compared to the LR model with EHR data (magenta trace—Figure 3A and 3B) with mean ROC AUC 0.83 +/- 0.01 and PR AUC of 0.33 +/- 0.03).

#### Prospective validation on Device #1 data

Using the prospective dataset held out entirely for testing from Device #1 (separate hospital), the model performance on this retrospective dataset (blue trace—Figure 3C and 3D) had ROC AUC 0.9 and PR AUC 0.6, with LR model showing worse performance (ROC AUC 0.83 and PR AUC 0.3, green trace—Figure 3C and 3D and Table 2). The RNN-based model using EHR data (turquoise trace—Figure 3C and 3D) alone showed a degradation in performance with ROC AUC 0.82 and PR AUC 0.49 and LR model with EHR data (magenta trace—Figure 3C and 3D) having ROC AUC 0.89 and PR AUC 0.43.

#### Alternate device testing on Device #2

All data from Alternate Device #2 was held out for testing and used for external validation of the RNN-based model. The model performance showed (blue trace—Fig. 5E and 5F) ROC AUC 0.84 and PR AUC 0.37 (LR model ROC AUC 0.85 and PR AUC 0.09, green trace—Figure 3E and 3F and Table 2). Using EHR data to predict alerts, the RNN-based model (turquoise trace, Figure 3E and 3F) had ROC AUC 0.82 and PR AUC 0.4 (LR with EHR model; magenta trace— Figure 3E and 3F, ROC AUC 0.89 and PR AUC 0.14).

Confusion matrix for each of the three validation steps (Device #1 retrospective, Device #1 prospective, Device #2 alternate). The operating point was chosen such that the PPV was equal to 0.2. The threshold that achieved this PPV is displayed in the upper left corner of each table section. Summary metrics (i.e., specificity, sensitivity, PPV, and negative predictive value) are listed at the periphery of each table.

### Testing RNN and LR model accuracy on hard outcomes

Using the RNN model trained on detecting > 30-minute periods of elevated MEWS, we tested the model’s performance on predicting hard outcomes within 24 hours (Table 3). In summary, 50% of RRT calls, 83.6% of unplanned transfers to the ICU, 100% of intubation/mechanical ventilation, 100% of cardiac arrests, and 100% of death were detected using CM data. This performance was superior to that of using solely EHR data or an elevated MEWS as the predictor for a hard outcome.

**Table 3:** Summary of RNN vs MEWS performances on hard outcomes using both wearable and EHR data.

|  | Wearable RNN<br>accuracy<br>(Correct/Total) | EHR RNN accuracy<br>(Correct/Total) | Wearable MEWS<br>accuracy<br>(Correct/Total) | EHR MEWS<br>accuracy<br>(Correct/Total) |
| --- | --- | --- | --- | --- |
| RRT | 50.0 (1/2) | 100.0 (2/2) | 0.0 (0/2) | 0.0 (0/2) |
| ICU Transfer | 83.3 (5/6) | 50.0 (3/6) | 16.7 (1/6) | 0.0 (0/6) |
| Ventilation | 100.0 (1/1) | 100.0 (1/1) | 0.0 (0/1) | 0.0 (0/1) |
| Cardiac Arrest | 100.0 (1/1) | 0.0 (0/1) | 0.0 (0/1) | 0.0 (0/1) |
| Deaths | 100.0 (1/1) | 0.0 (0/1) | 0.0 (0/1) | 0.0 (0/1) |

### Lead time and sampling rate analysis

To characterize the timing of predictions relative to alerts, we examined lead times of predictions of the RNN model for each alert and hard outcome. Lead times are the first true positive preceding actual alerts and hard outcomes. We visualized the true positives on a raster plot that showed all RNN-based alerts 24 hours preceding a clinical alert (Figure 3G) along with the distribution of lead times (Figure 3H). For the three stages of validation—Device #1 retrospective, Device #1 prospective, Device #2 alternate—the median and interquartile range (IRQ) of lead time was 17.12 [6.81 – 24.0], 24.0 [22.88 – 24.0] and 8.25 [5.25 – 21.75], respectively. For this analysis, we limited the scope of true positives to 24 hours, though several alerts for each validation had lead times greater than 24 hours. The percentage of alerts whose lead time was greater than 24 hours for Device #1 retrospective, Device #1 prospective, Device #2 alternate was 14%, 75%, and 21%, respectively.

We repeated this analysis for hard outcome predictions with all true positives preceding an outcome being visualized (Figure 3I). The timing of these first true positives is plotted in a probability density histogram plot with median (IQR) lead time of 16.5 [10.5 – 17.5] (Figure 3J).

To determine the effect of sampling rate on prediction performance, we varied the inter-sample timing of vital measurements from one to three hundred minutes and measured the effect on ROC AUC and PR AUC at each inter-sample interval (Supplementary Figure 3). Going from one to three hundred minutes, the ROC AUC dropped 16.2% and the PR AUC dropped 17.0%. The ROC and PR AUC maximum occurs at inter-sample intervals of one and thirty minutes, respectively.

## Discussion

In this study, we sought to investigate three lines of inquiry with respect to a wearable biosensor–based clinical alert system: (1) are the wearable devices comparable to manual episodic vital sign measurements, (2) do they provide an advantage over episodic monitoring in producing more timely and earlier clinical alerts, and (3) can wearable biosensor data be used in a deterioration prediction algorithm. With CM data on 888 patients across four hospitals, we found that CM- and EHR-based data are congruent and that, primarily due to high frequency, accurate RR, and HR measurements, wearable devices generate more and earlier clinical alerts. Finally, we showed that a deep-learning model based upon CM data accurately predicts patient deterioration up to eight to twenty-four hours before a MEWS-based clinical alert and seventeen hours before an actual clinical event.

The current standard of care in patients admitted on medical and surgical floors, where the study took place, implements a vital sign check every 3-4 hours depending on the hospital and unit where a patient is situated. When a patient reaches a MEWS score above seven, vital sign frequency is then increased to every 2 hours. A MEWS score of eight results in increased vital sign frequency and an independent provider evaluation. Supplementary Table 3 details the additional steps that are taken as the MEWS score increases beyond eight. In this study, we only looked at MEWS alerts (MEWS > 6 for 30 minutes or longer) that were not preceded by any other MEWS alerts for at least 24 hours. Therefore, the additional, earlier clinical alerts that were found in this study, represent truly unexpected deteriorations enabled by more frequent monitoring of the patient.

To address the concern of a selection bias existing in our patient selection process, we’d like to point out our deterioration event rate of ∼1% was actually lower than what is seen in the clinical literature (∼6%)^1^. While clinicians were basing the patch decision on their best clinical judgement, this didn’t lead to an increased deterioration event rate, for a variety of reasons, either due to: the acuity of the wards, the patching occurring outside of acute time periods, or nurses patching patients based on assessment of risk while also waiting until a convenient time to patch them. If a selection bias existed that benefitted the model, i.e. more deterioration event labels and more balanced data for the model to train more effectively on, we would expect to see higher rates of deterioration, which we don’t see. The patching scheme thus reflected real world conditions and therefore a real world scenario of the patching situation.

Several studies have validated CM devices in real-world settings and found that these devices are comparable to current gold standard of vital measurements^46–48^. Using common evaluation methods, we show accuracy results consistent with previously published results. While a formal measurement validation study is usually carried out in controlled settings with matched controls, this validation step was not the primary focus of our study. However, it provides evidence on these devices’ performance in real-world conditions—i.e., in a hospital environment with rotating clinical staff—and is an important part of our proposed framework. It should be noted that, while certain vital signals had an error magnitude that might preclude it’s use in clinical decision making (e.g. temperature), the goal in this paper was to capture trends via continuous monitoring wearable devices to predict adverse clinical events. It was not our intention in this paper to try to justify the use of raw vital signal readings from clinical wearables for use in clinical decision making. Since the vitals that went into the predictive algorithm were normalized, the absolute error of each vital signal was therefore not as important as the trends.

In our analysis, we attempted to rule out confounders, such as patient movement, in producing false alerts from CM data. We found no correlation between increased movement and occurrence of alerts or elevated vitals. Thus, the additional alerts we report are due to real vital signal aberrations caused by physiological decline or deterioration. We also investigated the driving components that led to these additional wearable-based alerts and found that respiratory rate and heart rate were the two major factors. This is not a surprising result for RR, since inaccurate documentation of RR in the EHR is well known due to digit preference and the increased burden of measuring and reporting an accurate RR^49^. On the other hand, while measured accurately in the EHR, HR can provide more insights when measured more frequently (likely because of its highly dynamic nature). It should be noted however, that this study was not intended to comment on the clinical utility of the additional wearable generated alerts, since all of the additional alerts were generated post-hoc and offline.

Our results show that the proposed deep learning model performs better with CM data than with episodic monitoring data. In addition to the increased accuracy of RR and HR CM measurements (shown in several studies to be two of the most important factors in predicting risk of deterioration), this outcome is due to the temporally richer nature of the CM data that better represents subtle signs of deterioration^26,50–52^. However, even when CM devices are unavailable, the performance of the algorithm using only episodic EHR data remains comparable to that of other deterioration prediction algorithms^23,25,27,32,41,53–55^ thus demonstrating the versatility of the proposed approach.

We’d also like to note the higher, and comparable, AUROCs for the Device #2, and Device #1, logistic regression EHR models, respectively. These anomalies are a good example of why the inclusion of the Precision-Recall curve is important—the ROC plot can be misleading on the performance of a classifier on a highly imbalanced dataset such as the one in this study (99.7% / 0.3% negative / positive class balance). Given this, we’d like to point out that the RNN significantly outperformed the logistic regression model on the precision-recall curve for the Prospective Device # 1 validation and both EHR and wearable data being comparable for the Device # 2 validation. This discrepancy captures the fact that the logistic regression model simply performed better than the RNN on predicting true negatives (hence the higher AU-ROC), however the RNN was still significantly superior on predicting true positives (hence the higher AU-PR).

In our analysis, we designated false negatives as more detrimental than false positives because the main goal of our algorithm is to identify patients experiencing unanticipated deterioration, i.e. minimizing missed deteriorations was the priority. This approach could potentially lead to an increased number of alerts. To avoid contributing to alert fatigue, a balanced choice of a minimum positive predictive value needs to be made. In our study, a tentative choice of positive predictive value (PPV) = 0.2 (i.e., 1 true positive per 5 alerts) was made; this is a stricter value than the usual 0.1 PPV used in prior studies^56^. However, one of the benefits of our approach is that the alerting threshold can be adjusted based upon risk tolerance and resource considerations.

To assess model calibration, we looked at calibration curves of the LSTM RNN model for each stage of validation. The results are not surprising, given that modern neural network architectures are known to exhibit overconfidence, pushing predicted probabilities toward 0 or 1, mostly arising from the interplay between softmax/sigmoid activations and unregularized logit magnitudes. This “confidence sharpening” is evident in our Retrospective Device #1 calibration curve (“Device #1 Retrospective” Supplementary Figure. 5). Between approximately a 5% and 10% fraction of positives, the mean predicted probability jumps from 10% to >95%, since the model has set its decision boundary at a point in the input space corresponding to approximately a 7% true risk of deterioration (i.e. fraction of positives) and scales the risk exponentially as one crosses this boundary. The same behavior is seen in the prospective validation calibration curve, however since the dataset is smaller and there are less acute patients, the highest risk patients in this dataset are only at a ∼10% risk of deterioration as opposed to the ∼25% seen in the retrospective dataset. The Alternate Device #2 dataset takes this point even further in that the highest risk group of patients only have ∼1-2% chance of deterioration. Additionally, given the multiple nested non-linear activation functions inherent in a deep neural network LSTM architecture, it is not surprising that this produces a non-continuous, non-linear numerical output that does not correspond to an estimate of the risk of deterioration. While it is evident from the calibration curve and Brier scores that the model is not well initially well calibrated, this does not undermine our findings in the ROC and PR of the model’s strong discriminatory ability. Finally, recalibration can correct model calibration across the observed ranges of patient risk.

We also showed preliminary capability of the model in predicting a broad class of “hard outcomes” against which the model was not trained. Despite having very few of these outcomes, the algorithm performed quite well in detecting a broad class of them and demonstrates the utility of our approach in using clinical alerts as a proxy label for model training. We also demonstrate the advantage of an RNN-based algorithm with CM data, as it outperformed a MEWS-based alert and alerts using EHR data alone. Further, by looking at the average lead time of the model predictions with respect to hard outcomes, we show that risk for these outcomes could be detected between 12 to 18 hours in advance and allow for anticipatory clinical intervention.

The lead time analysis of our predictive model revealed that we can expect to predict clinical alerts up to eight to twenty-four hours in advance and hard outcomes up to seventeen hours in advance, thus hospital staff will have many opportunities to intervene and change the trajectory a deteriorating patient’s physiological decline. This intervention could look much like the current protocol in place for existing early warning score systems (Supplementary table 3) except with further integration into wearable dashboards in addition to smarter EHR alerting. Moreover, our approach offers other implementation possibilities—e.g., the predictions could be served to a central monitoring team, displayed to clinicians in the EHR, or sent to a clinician as a direct notification via a secure text message.

By downsampling the continuous vital signals and measuring the impact, we found that a lower sampling rate does not degrade the predictive performance of the algorithm and, in fact, may slightly improve it at an inter-sample interval of 30 minutes. Notably, we did not train the RNN model with downsampled data, and a higher sampling rate may be needed initially to train models with subsequent deployment using lower sampling rates. If true, this lower sampling rate could extend the battery life and overall longevity of single use devices, including those used in this study, thus lowering costs for hospitals to implement continuous monitoring via wearable devices as a standard of care for non-ICU inpatients. This contention, however, requires further study and evaluation.

This study had several limitations. It did not implement eligibility criteria during participant selection that would maximize focus on patients that experience adverse events more frequently. For this reason, the total number of hard outcomes was under 2% (i.e., 11 total outcomes). While this small number made training a model specifically for said outcomes infeasible, our limited validation of the deep learning algorithm on these events showcases that this approach could be applicable to prediction of hard outcomes. It should be noted that, while the limited validation on the hard outcomes is promising, the small number of events just points to feasibility of developing a model capable predicting hard outcomes. However, no strong conclusions can be made. Collecting more data with more patients and more hard clinical outcomes will enable a better validation of this algorithm on hard outcome prediction capabilities. Stricter eligibility criteria for outfitting patients with CM patches, including MEWS thresholds upon hospitalization or focus on higher-risk comorbidities/diagnoses, could maximize the number of hard outcomes captured while patients are monitored continuously, enabling better training and validation of future models on such hard clinical outcomes.

## Conclusion

This study presents a framework that uses continuously monitored vital signs obtained via wearable devices to predict clinical alerts related to inpatient deterioration. Through validation and subsequent analysis of the continuously measured vitals against episodically measured vitals and the resulting clinical deterioration alerts, we show that use of CM data can result in more and earlier clinical alerts compared to EHR data alone. We also present a deep learning algorithm that uses CM and EHR data to predict these clinical alerts up to 24 hours ahead of time, which maintains performance across different patient cohorts, hospital settings, and CM devices. The algorithm shows promise in predicting hard outcomes, including ICU transfer and death. To our knowledge, this is the first study of its size to propose a comprehensive inpatient CM framework tested on hard clinical outcomes that is validated against real-time deployment scenarios.

## Acknowledgements

We thank Andrew Dominello and Jennifer Connell for their help in the copyediting and submission of this article. This work is supported by NIH 5R01NR020774-02.

## Data Availability

The data that support the findings of this study contain protected health information (PHI) and cannot be completely deidentified, due to the importance of timestamps on both the continuous monitoring data, and the EHR variables in our analysis. They cannot be shared publicly due to privacy and confidentiality concerns and access to the data is restricted in accordance with patient privacy regulations and institutional policies. Researchers who meet the criteria for access to confidential data may submit a request to the Northwell Institutional Review Board (IRB) for consideration. For inquiries regarding data access, please contact the IRB at.

## Code Availability

The underlying code for this study is not publicly available but may be made available to qualified researchers on reasonable request from the corresponding author.

## Supplementary Materials

**Supplementary Table 1.** Cutoff values for various classes of movement derived from Device #1’s signal magnitude area (SMA) signal which is the average output of a 3-axis accelerometer.

| Percentiles | SMA values (mg/s) |  |  |
| --- | --- | --- | --- |
|  | Stationary | ADL | Strenuous |
| 25 | 8 | 10 | 94 |
| 50 | 8 | 26 | 262 |
| 75 | 10 | 118 | 456 |

**Supplementary Table 2.**
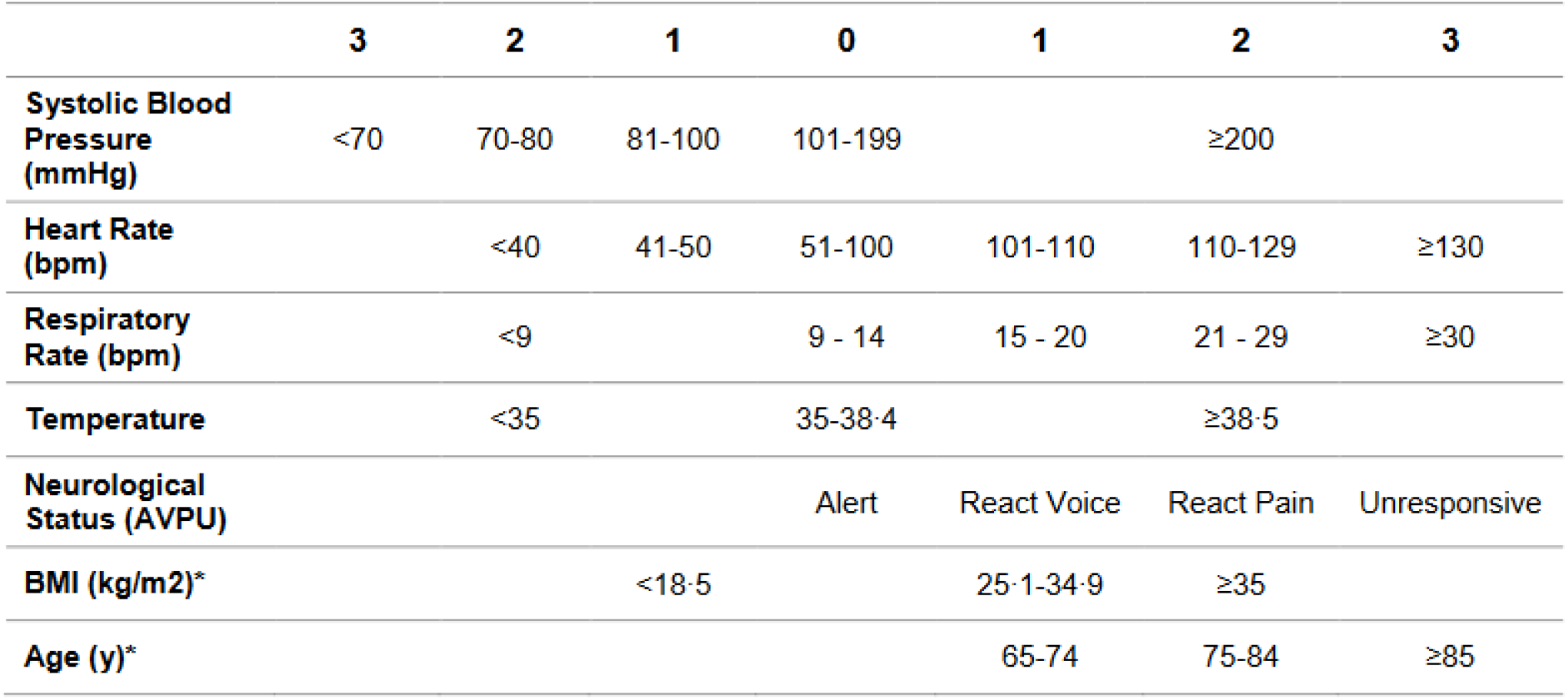
Modified Early Warning Score (MEWS) calculation, based upon vital sign measurements. Each item is given a score, and the final score is tallied (range 0-15), with higher values indicating greater risk of clinical decompensation.

**Supplementary Table 3.**
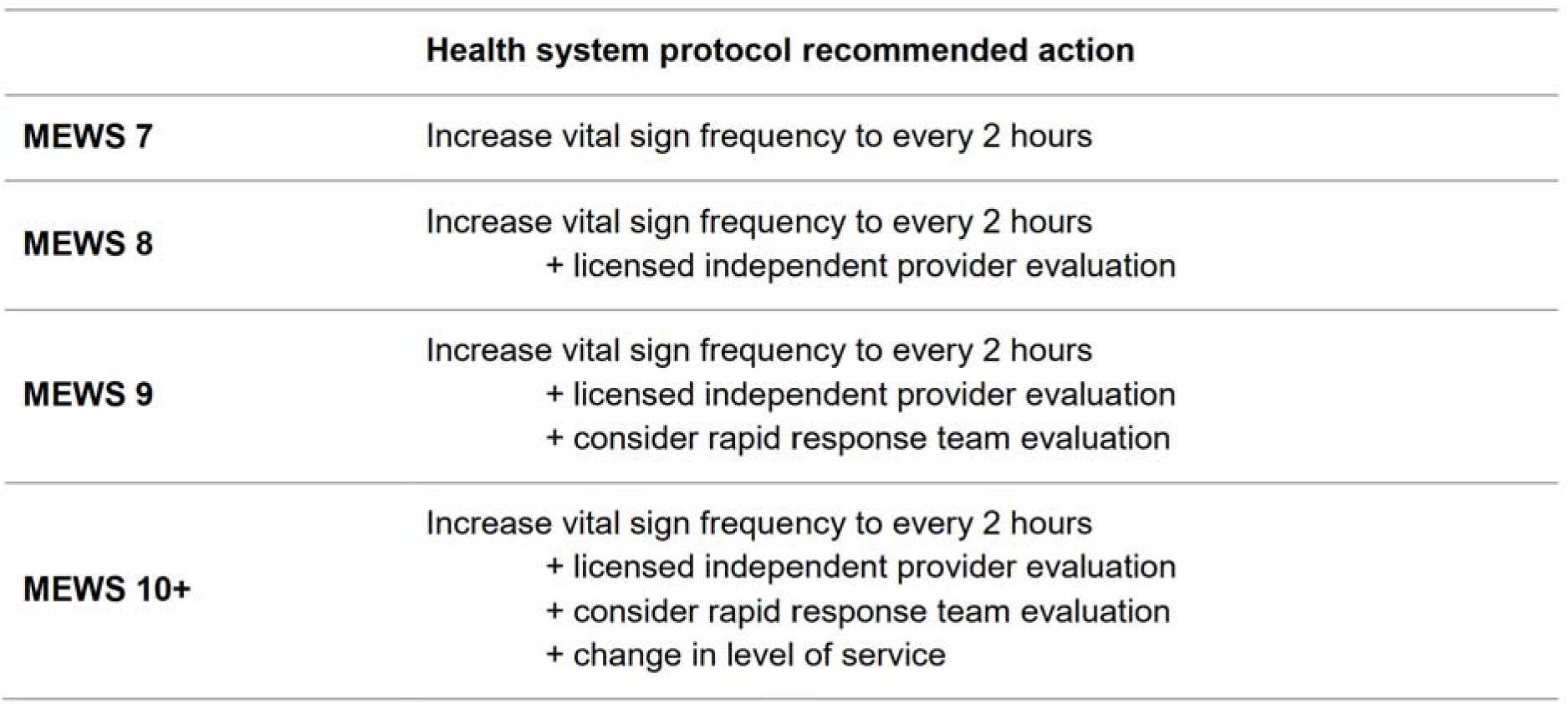
Health system protocol as defined by the Modified Early Warning Score (MEWS) value, incorporated into the electronic health record.

|  | Health system protocol recommended action |
| --- | --- |
| **MEWS 7** | Increase vital sign frequency to every 2 hours |
| **MEWS 8** | Increase vital sign frequency to every 2 hours<br>+ licensed independent provider evaluation |
| **MEWS 9** | Increase vital sign frequency to every 2 hours<br>+ licensed independent provider evaluation<br>+ consider rapid response team evaluation |
| **MEWS 10+** | Increase vital sign frequency to every 2 hours<br>+ licensed independent provider evaluation<br>+ consider rapid response team evaluation<br>+ change in level of service |

**Supplementary Table 4.**
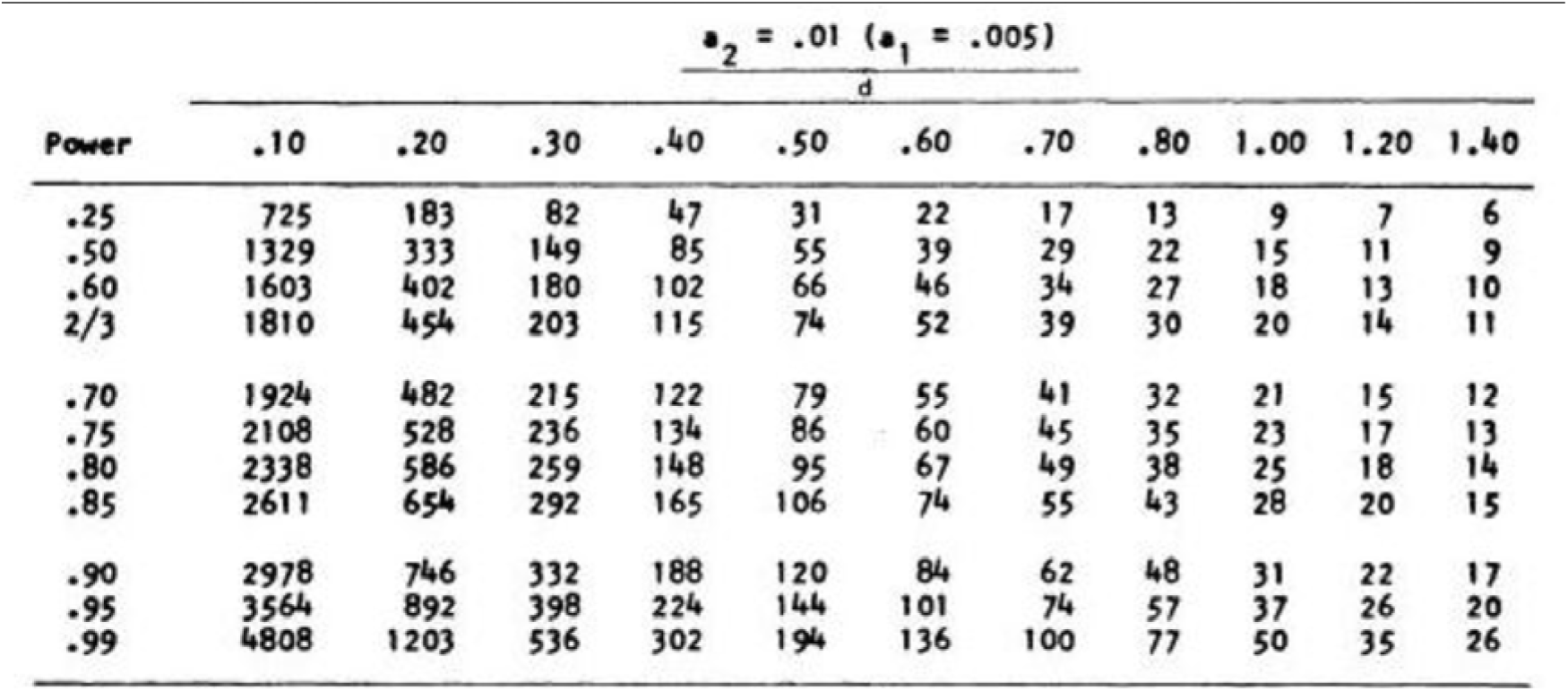
Sample size estimation for wearable data validation and model convergence. To estimate the sample size necessary for validating the wearable data in this study, we calculated Cohen’s d statistic using the following formula:

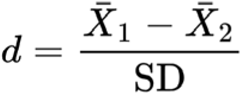 Where _1_ is the mean of the distribution of vital values measured from the wearable device and _2_ is the mean of the distribution of vital values measured in the EHR. SD represents the expected standard deviation of the grouped data. Using this formula, we set out to test whether the difference in the means of the wearable and EHR vital values exceeded 10%, in other words, whether there was a 10% error or greater. To give a concrete example, assuming an expected average heart rate of 75bpm, a 10% difference in the means of the heart rates, from wearable and EHR sources, would translate to a difference of the means of at least 7.5bpm. The expected aggregate standard deviation of the heart rate from both sources is approximately 10 bpm. Entering these values into the equation above results in a Cohen’s d of 0.75. For this study, we chose a power level of 95% with alpha = 0.01. Using these parameters and the following sample size lookup table: ^62^ Resulted in an estimated required sample size between 57 and 74. Since each of our participants had many measurements, our study far exceeded the required sample size to detect the error rate in vital sign measurement between the wearable device and EHR. In addition to estimating the sample size for validating the wearable data, we also estimated the amount of data necessary for neural network convergence. The choice of cohort size was made based on other clinical studies with similar architectures,^59, 60^ our prior studies, ^61^ as well as studies on VC dimensions of neural networks.^58^ With this context in mind, we observed that LSTM RNNs typically converge on sample sizes with over 600 patients. Recruiting 888 patients was based on this consideration and the availability of patches during this study.

**Supplementary Table 5.** Missing data from CM devices from preliminary data from 888 patients.

| Missingness (%) | VitalPatch | Biobeat |
| --- | --- | --- |
| Respiratory Rate | 3.72 | 8.3 |
| Heart Rate | 3.72 | 0 |
| Body Temp. | 0 | 3.9 |
| SpO2 | 14.95 | 23.02 |
| Activity (SMA) | 0 | 0 |

**Supplementary Figure 1.**
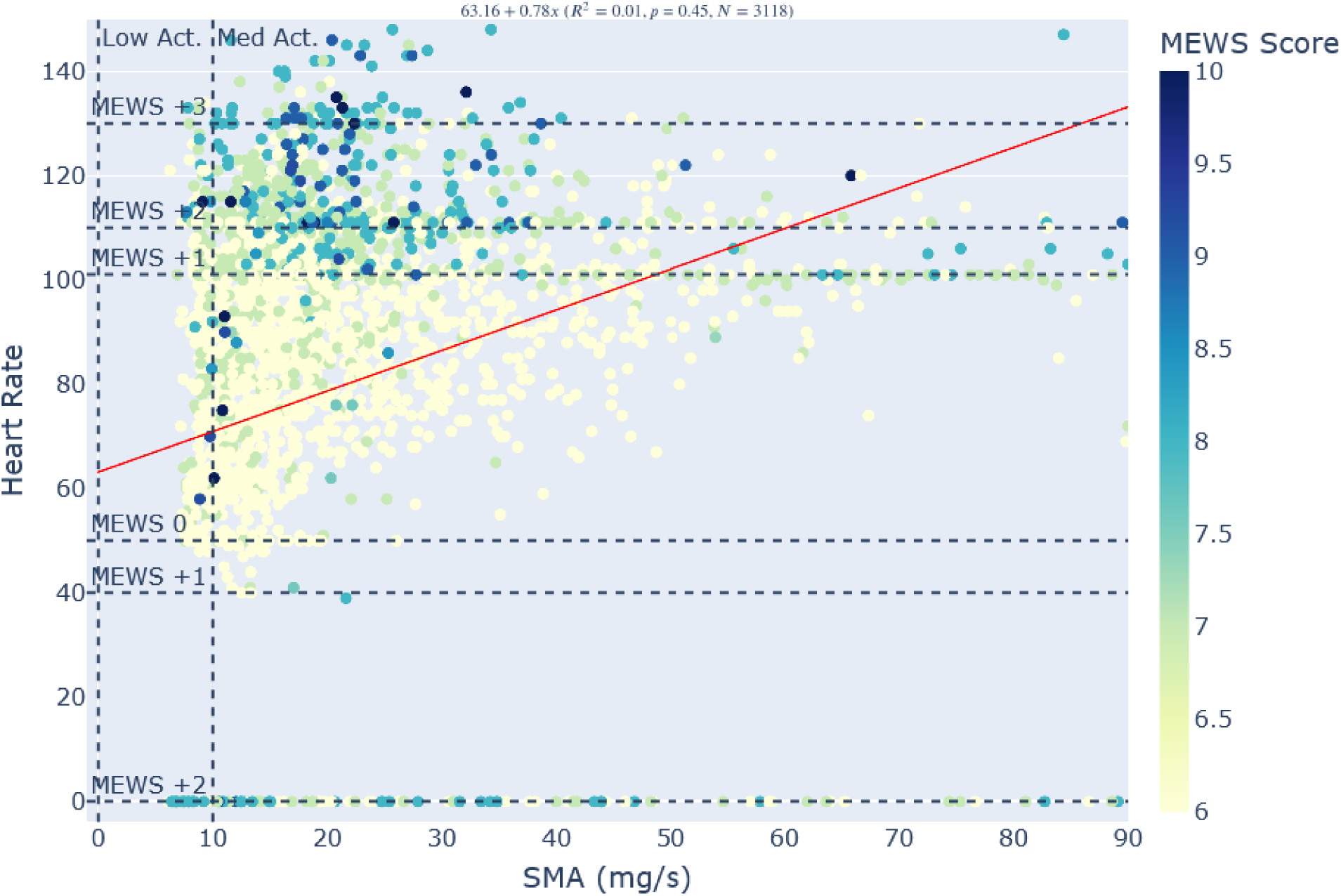
shows the relationship between signal magnitude area (SMA) and heart rate. To test whether movement (SMA) may be contributing to vital sign derangement and thus elevated MEWS values, we examined the values of HR against the average SMA values over the duration of the detected alert and associated MEWS value. The alerts and their corresponding average SMA values, were grouped into two classes (vertical dotted lines): “stationary” or Low Activity (Low Act.) and “activities of daily living” or Medium Activity (Med. Act.), based on manufacturer determined SMA cutoff values (Supplementary Table 1). The heart rate was then also grouped into six classes depending on its point contribution (Supplementary Table 2) to the composite MEWS score (horizontal dotted lines). Each SMA-HR pair was then colored by the average MEWS score over the duration that the MEWS score was elevated for that alert. The linear fit equation, correlation coefficient and significance sit at the top of the figure.

**Supplementary Figure 2.**
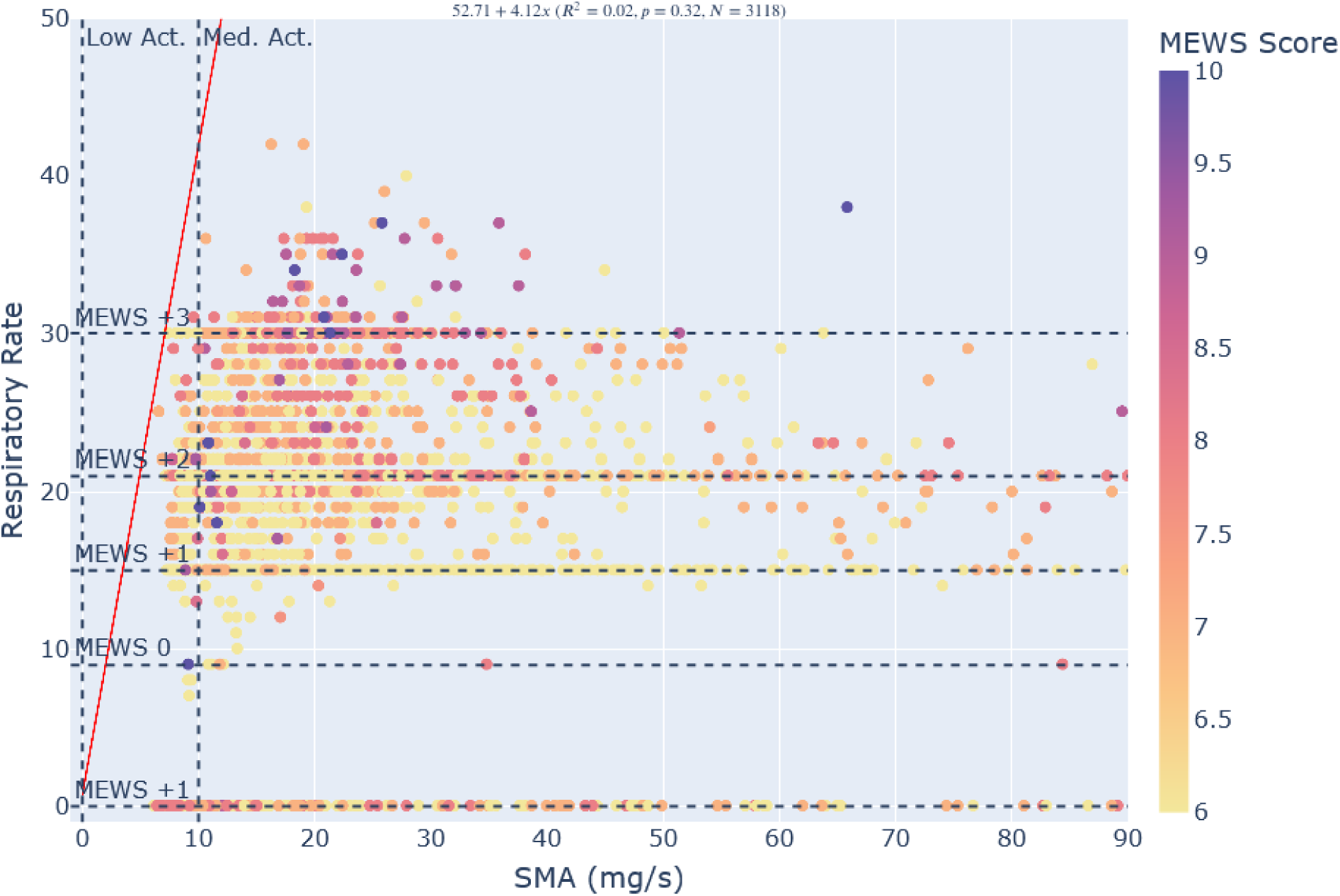
shows the relationship between movement (SMA) and respiratory rate (RR). To test whether movement (SMA) may be contributing to vital sign derangement and thus elevated MEWS values, we examined the values of RR against the average SMA values over the duration of the detected alert and associated MEWS value. The alerts and their corresponding average SMA values, were grouped into two classes (vertical dotted lines): “stationary” or Low Activity (Low Act.) and “activities of daily living” or Medium Activity (Med. Act.), based on manufacturer determined SMA cutoff values (Supplementary Table 1). The RR was then also grouped into five classes depending on its point contribution (Supplementary Table 2) to the composite MEWS score (horizontal dotted lines). Each SMA-RR pair was then colored by the average MEWS score over the duration that the MEWS score was elevated for that alert. The linear fit equation, correlation coefficient and significance sit at the top of the figure.

**Supplementary Figure 3.**
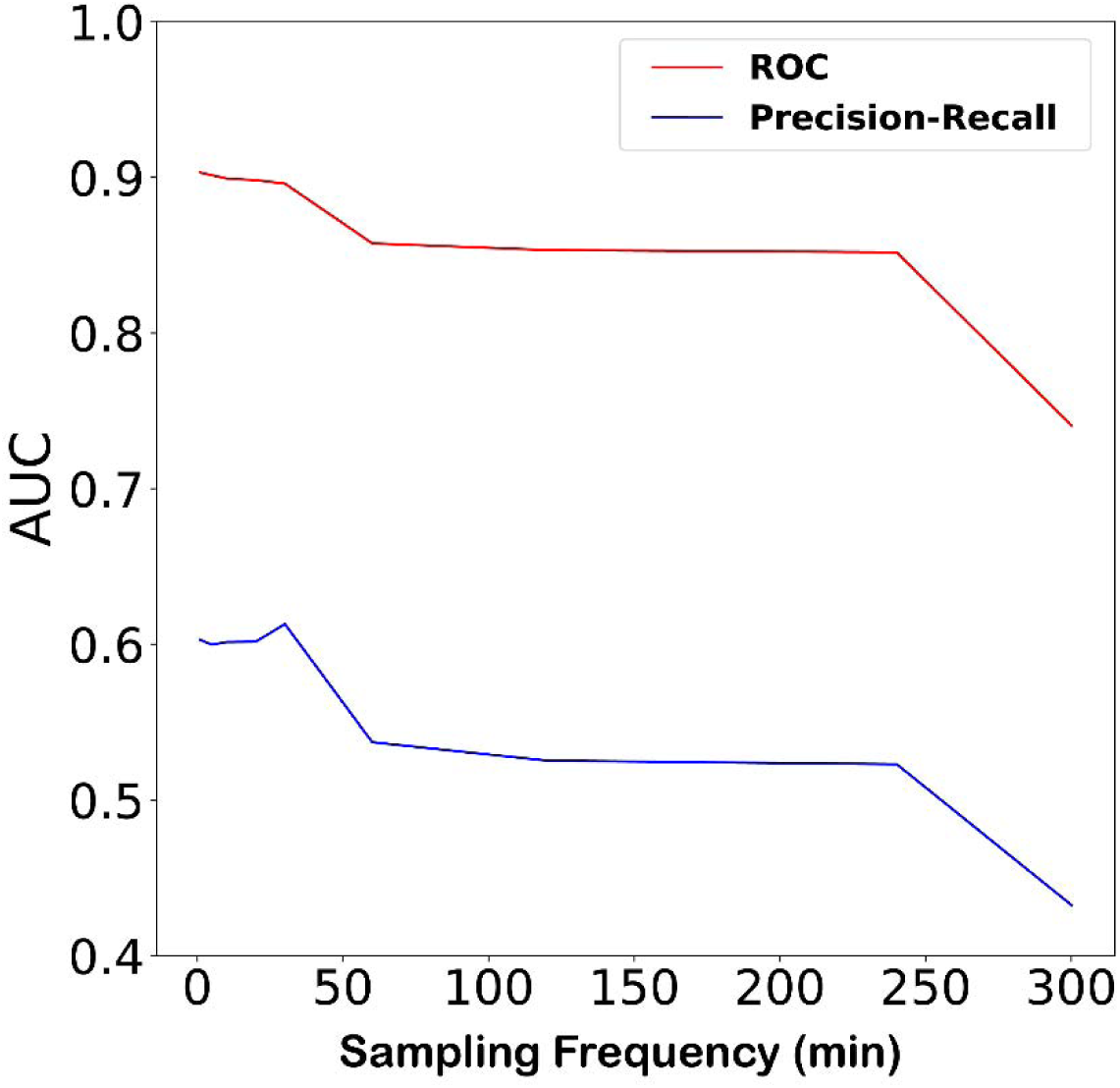
Relationship between inter-sampling interval and performance of the wearable based RNN deterioration prediction algorithm. ROC AUC (red trace) and PR AUC (blue trace) metrics show the effect of inter-sample interval on prediction performance as the inter-sample interval of vital measurements is extended out from 1-minute to 300minutes.

**Supplementary Figure 4.**
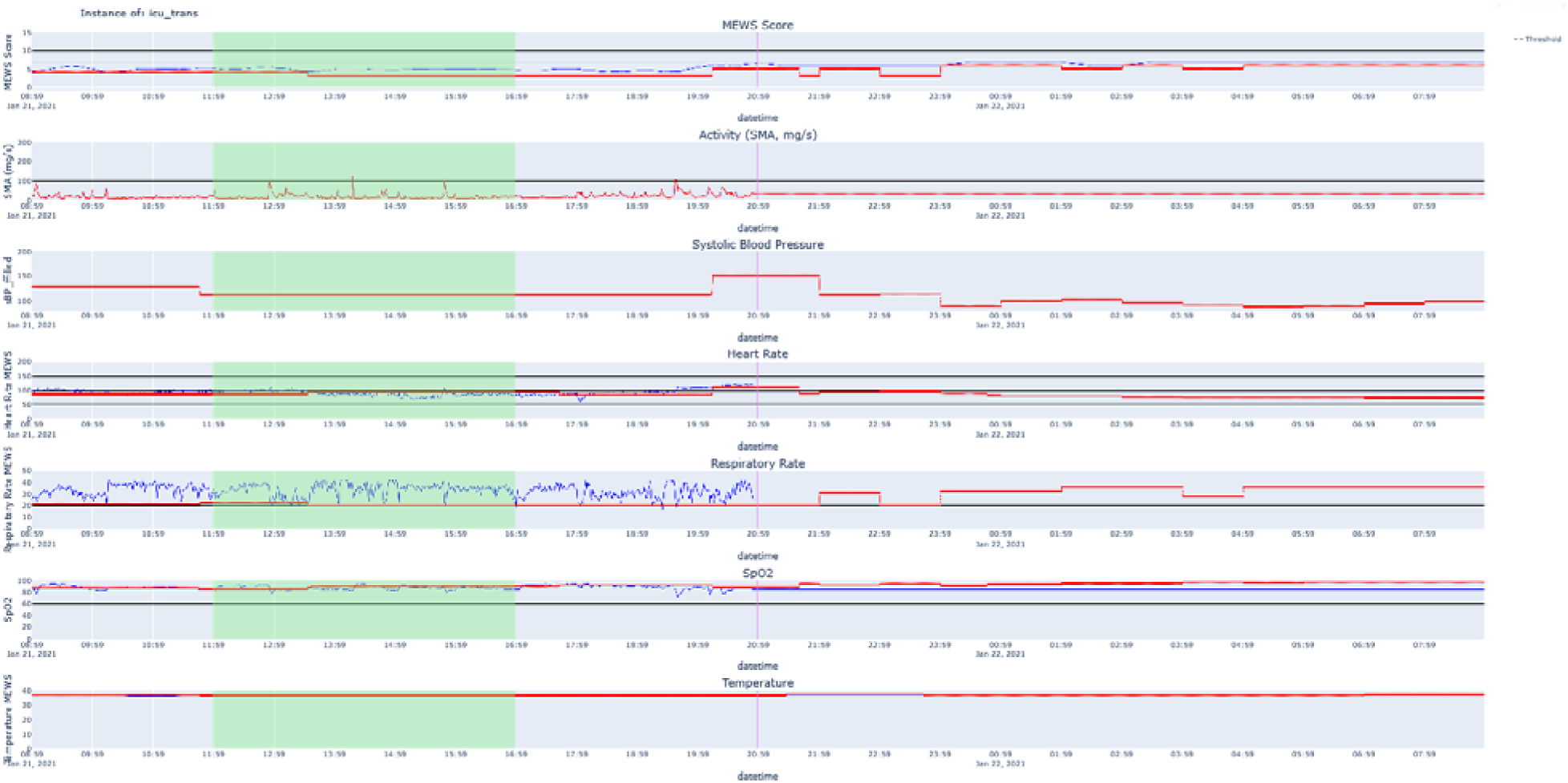
Sample patient vitals preceding an ICU transfer event. Sample patient vitals preceding an ICU transfer event are shown of: manually measured vital values input by a nurse into the EHR (red traces), the most recently calculated MEWS score based on all of the manually measured vitals in the EHR (red trace, top plot), the most recently measured wearable value for that particular vital (blue traces), and the most recently calculated MEWS score based on the continuously measured vital signals from the wearable device (blue traces, top plot). The black dotted line in the top plot represents the MEWS 6 threshold that was used for identifying clinical alerts. The vertical magenta line indicates the time of the ICU transfer event. The green shaded area represents an example 5-hour sequence of data that is extracted and passed into a model to try to predict the ICU transfer event, in this case with a three-hour lead time.

**Supplementary Figure 5.**
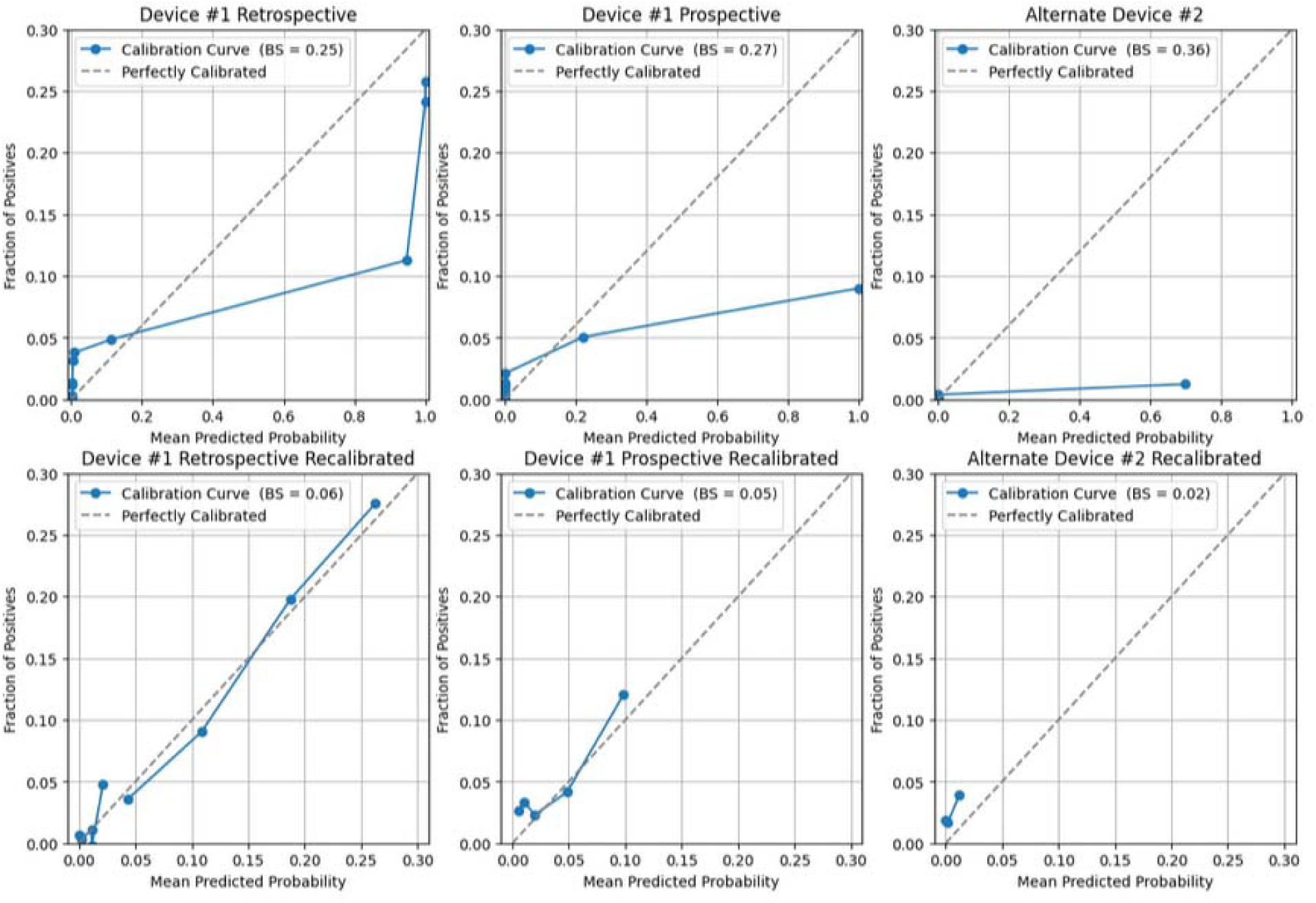
Calibration Curves for the LSTM RNN Model. Calibration curves (blue traces) across the three stages of validation Device #1 Retrospective (left), Device #1 Prospective (center) and Alternate Device #2 (right). The first row shows original calibration curves for each stage of validation from left to right. The second row shows the recalibrated calibration curves for each stage of validation from left to right. Brier scores (BS) are included in the figure legend. Perfect calibration (dotted line) is shown in each case.

